# Using Supervised Machine Learning and Empirical Bayesian Kriging to reveal Correlates and Patterns of COVID-19 Disease outbreak in sub-Saharan Africa: Exploratory Data Analysis

**DOI:** 10.1101/2020.04.27.20082057

**Authors:** Amobi Andrew Onovo, Akinyemi Atobatele, Abiye Kalaiwo, Christopher Obanubi, Ezekiel James, Pamela Gado, Gertrude Odezugo, Dolapo Ogundehin, Doreen Magaji, Michele Russell

## Abstract

**Introduction:** Coronavirus disease 2019 (COVID-19) is an emerging infectious disease that was first reported in Wuhan^1,2^, China, and has subsequently spread worldwide. Knowledge of coronavirus-related risk factors can help countries build more systematic and successful responses to COVID-19 disease outbreak. Here we used Supervised Machine Learning and Empirical Bayesian Kriging (EBK) techniques to reveal correlates and patterns of COVID-19 Disease outbreak in sub-Saharan Africa (SSA).

**Methods:** We analyzed time series aggregate data compiled by Johns Hopkins University on the outbreak of COVID-19 disease across SSA. COVID-19 data was merged with additional data on socio-demographic and health indicator survey data for 39 of SSA’s 48 countries that reported confirmed cases and deaths from coronavirus between February 28, 2020 through March 26, 2020. We used supervised machine learning algorithm, Lasso for variable selection and statistical inference. EBK was used to also create a raster estimating the spatial distribution of COVID-19 disease outbreak.

**Results:** The lasso Cross-fit partialing out predictive model ascertained seven variables significantly associated with the risk of coronavirus infection (i.e. new HIV infections among pediatric, adolescent, and middle-aged adult PLHIV, time (days), pneumococcal conjugate-based vaccine, incidence of malaria and diarrhea treatment). Our study indicates, the doubling time in new coronavirus cases was 3 days. The steady three-day decrease in coronavirus outbreak rate of change (ROC) from 37% on March 23, 2020 to 23% on March 26, 2020 indicates the positive impact of countries’ steps to stymie the outbreak. The interpolated maps show that coronavirus is rising every day and appears to be severely confined in South Africa. In the West African region (i.e. Burkina Faso, Ghana, Senegal, Cote d’Iviore, Cameroon, and Nigeria), we predict that new cases and deaths from the virus are most likely to increase.

**Interpretation:** Integrated and efficiently delivered interventions to reduce HIV, pneumonia, malaria and diarrhea, are essential to accelerating global health efforts. Scaling up screening and increasing COVID-19 testing capacity across SSA countries can help provide better understanding on how the pandemic is progressing and possibly ensure a sustained decline in the ROC of coronavirus outbreak.

**Funding:** Authors were wholly responsible for the costs of data collation and analysis.

## Introduction

On Jan 30, 2020, WHO declared the current novel coronavirus disease 2019 epidemic a Public Health Emergency of International Concern^3^. As of March 26, 2020, the number of COVID-19 cases surpassed 500,000 globally, and the epidemic registered a total of 1,978 cases and 25 deaths in sub-Saharan Africa (SSA). At this time, 39 of SSA’s 48 countries [excluding Burundi, Comoros, Lesotho, Sierra Leone, South Sudan and Sao Tome and Principe] all reported confirmed cases of coronavirus and imported transmission as the major mode of spread. Despite aggressive response by SSA countries to stymie the outbreak, the number of reported new cases and deaths continue to increase. As of March 30, 2020, South Africa is the only country with more than 1,000 cases, and eight countries: Burkina Faso, Cameroon, Cote d’Ivoire, Senegal, Ghana, Mauritius and Nigeria including South Africa account for 80% of SSA’s COVID-19 disease outbreak (Figure 1).

**Figure 1:**
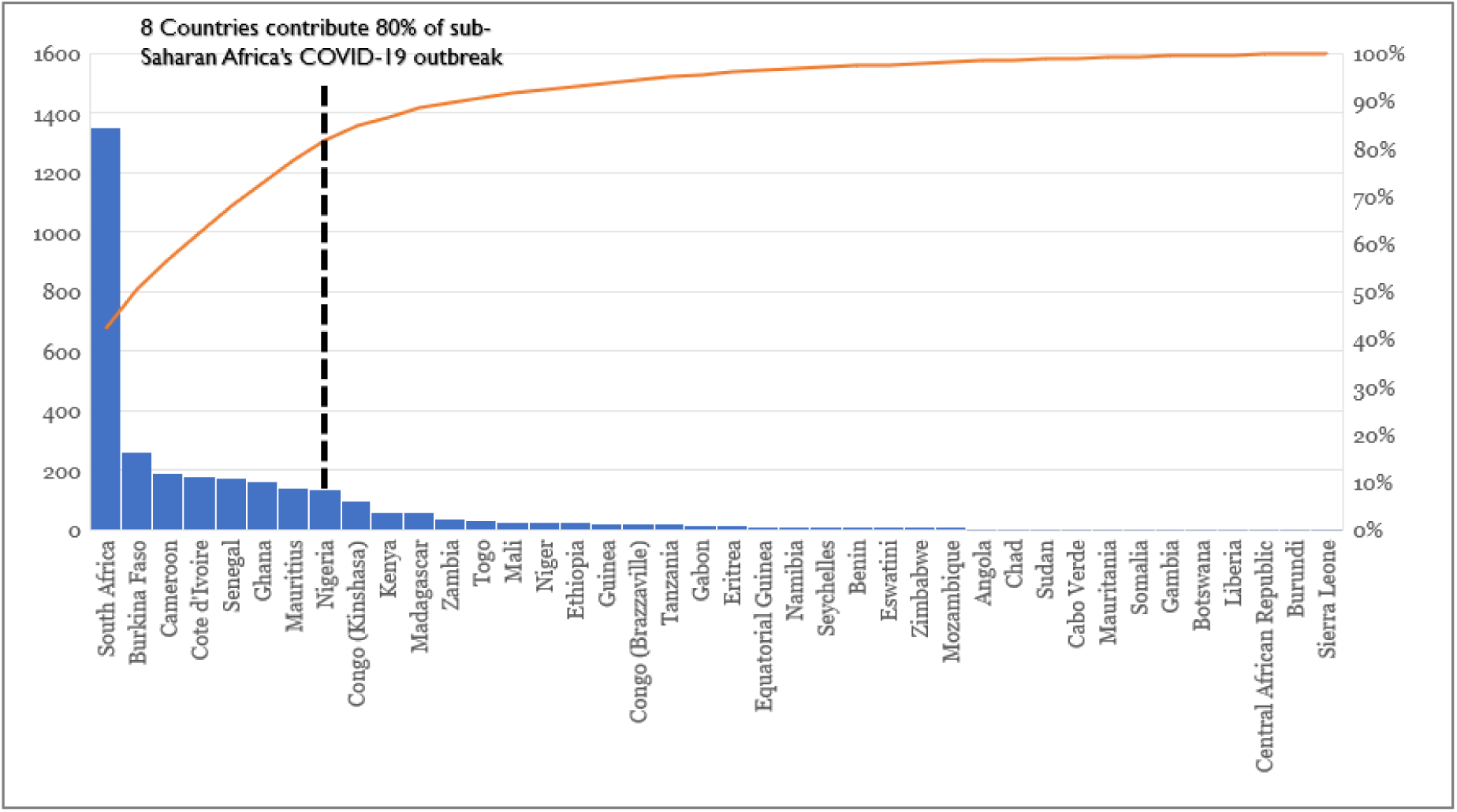
Countries contributing 80% of COVID-19 Disease Outbreak in SSA [February 28, 2020 through March 31, 2020].

As reported by Huang et al,^4^ patients with COVID-19 present primarily with fever, myalgia or fatigue, and dry cough. Although most patients are thought to have a favorable prognosis, older patients and those with chronic underlying conditions may have worse outcomes. Certain epidemiological features and clinical characteristics of COVID-19 have been previously reported^5,6^. However, these findings were based on prediction models or studies from America, Asia and Europe countries, and the use of these findings might be inappropriate for SSA context. The spatial distribution pattern of coronavirus disease outbreak and the risk factors leading to poor clinical outcomes have not been well understood and delineated.

In this study, we employed the exploratory data analysis (EDA) technique to maximize insights from our study data, uncover patterns and summarize risk factors associated with the coronavirus disease outbreak in SSA. EDA is an approach for data analysis that employs a variety of technique for summarizing and visualizing important characteristics of a dataset. This will be the first study to use supervised machine learning algorithm, “Least Absolute Selection Shrinkage Operator” (Lasso) regression as the principle method for variable selection (predict important variables associated with COVID-19) and for statistical inference (confirm or establish relationship of risk factors and COVID-19 disease); and Empirical Bayesian Kriging (EBK) used to create a raster estimating the temporal spatial distribution of COVID-19 disease outbreak in SSA.

## Methods

### Setting and study population

We collected and analyzed time series aggregate data on the 1,978 confirmed cases and 25 deaths of COVID-19 disease outbreak across SSA between February 28, 2020 through March 26, 2020. According to the United Nations, SSA consists of all African countries that are fully or partially located south of the Sahara. We focused on 48 of Africa’s 54 countries excluding Algeria, Djibouti, Egypt, Libya, Morocco and Tunisia, in accordance with the World Bank lists of countries in SSA.

### Data Sources

We accessed the COVID-19 Data Resource Hub developed by the Tableau community and utilized near-real time data compiled by Johns Hopkins University (JHU) and includes data from World Health Organization (WHO) and the Centers for Disease Control and Prevention (CDC). Additional data from socio-demographic and health indicator surveys was derived from resources of the World Bank, UNICEF, WHO and UNAIDS. This additional data was merged with the JHU COVID-19 data to establish the dataset that was used for this study. The dataset included the most recent and available data across SSA, and period of the data spanned between 2013 and 2019. For data on health systems performance index, these was derived from the WHO publication in 2001 entitled “Measuring Overall Health Systems Performance for 191 countries”.

### Measures

The target variable included in the supervised machine learning model was confirmed cases of Coronavirus disease. The target variable was log-transformed to control for skewness and ensure effective linear relationship with the independent variables. Explanatory or independent variables in the model included total population, GDP per capita, percentage of population with access to electricity, percentage of population with access to basic drinking water, incidence of malaria (per 1,000 population at risk), percentage of men and women aged 15 and over who currently smoke any tobacco product, Diarrhea treatment (percent of children under 5 receiving oral rehydration and continued feeding), percentage of infants who received third-dose of pneumococcal conjugate-based vaccine (PCV), incidence of tuberculosis (per 100,000 people), percent out-of-pocket expenditure, life expectancy at birth, Health Systems Performance Index, estimated incidence rate (new HIV infection per 1,000 uninfected population, children aged 0–14 years), estimated incidence rate (new HIV infection per 1,000 uninfected population, adolescents aged 10–19 years), HIV prevalence among people aged 15–49 years, transmission classification of COVID-19 disease (1=imported, 2=local transmission), income group (1=High Income, 2=Low income, 3=Lower middle income, 4=Upper middle income), Geocoordinates of SSA countries (latitude and longitude), and Time (days) between the first and last reported coronavirus cases. These variables were included in the analyses because recent research on coronavirus has found that biological variables such as pneumonia, malaria, and diarrhea^7^, socio-demographic variables like age^8^, access to comprehensive health care system^9^ were associated with the transmission of coronavirus. Consequently, other important variables particularly, cancer, diabetes or coronary heart disease (CHD) were not available for analysis.

## Analysis

### Machine Learning

We used Lasso regression a supervised machine learning method for 1.) prediction and model selection, and 2.) inferential statistics. For prediction and model selection, we used three separate selection models available in StataMP v.16. These models were Cross-validation (CV), Adaptive lasso and minimum Bayesian Information Criterion (BIC). In running Lasso, the dataset was split into two sample groups; Group 1 was training datasets we used to select our model, and Group 2 our testing datasets we used to test the prediction. We used Lasso because of its greater prediction accuracy and increased model interpretability. For the inferential statistics, we used the Cross-fit partialing out Lasso technique to estimate the coefficients, robust standard errors, p-values and confidence intervals of the specified variables of interest while the other covariates were selected as controls in the model.

### Geospatial Analysis

We georeferenced all confirmed cases and deaths from coronavirus that was reported across 39 countries and performed three analyses. First, we constructed maps of coronavirus infection across three different timepoints; time 1 (March 8, 2020), time 2 (March 14, 2020) and time 3 (March 26, 2020) and overlaid the coronavirus transmission classification type. Second, we used Spatial Autocorrelation (Global Moran’s I) statistics to measure the degree, to which coronavirus infection is clustered, dispersed or randomly distributed. The expected value under the null hypothesis is that *“there is no pattern of coronavirus infection in selected SSA countries”*. Moran’s I values range from −1 indicating perfect dispersion to +1 indicating perfect spatial clustering. A zero Moran’s I value indicates a random spatial distribution. Third, we used EBK technique in the Geostatistical Analyst tool of ArcGIS 10.8 software to estimate the distribution of coronavirus across SSA.

## Results

### Descriptive analysis

We analyzed 39 of SSA 48 countries that reported confirmed cases and deaths from coronavirus between February 28, 2020 through March 26, 2020. We plotted a matrix graph to explore the strength and association between 17 continuous variables and COVID-19. Overall, in figure 2a, the matrix graph showed that a majority, 82% (13/17) of the continuous variables indicated strong to moderate positive linear relationships, while two variables, Out-of-pocket expenditure and HIV incidence rate in children aged 0–14 years showed strong negative linear relationships. Prevalence of smoking was weakly correlated with coronavirus. Approximately 31% (12/39) of the SSA countries had missing information on smoking prevalence and this may have impacted on this variable as evident with the significant Chi-square goodness of fit *χ2* (1) = 5.77, *p*= 0.016. All other variables included in the analysis had complete data for almost all countries, however, in few countries where data was missing, this was ≤ 5%. In Figure 2b, Spearman’s rank-order correlation was run to determine the relationship between the two categorical variables and COVID-19. There was a moderate, positive correlation between transmission classification and COVID-19 disease, (*r_s_* (37) = 0.411, *p* = 0.009), with transmission classification explaining 17% of the variance in COVID-19 disease.

**Figure 2:**
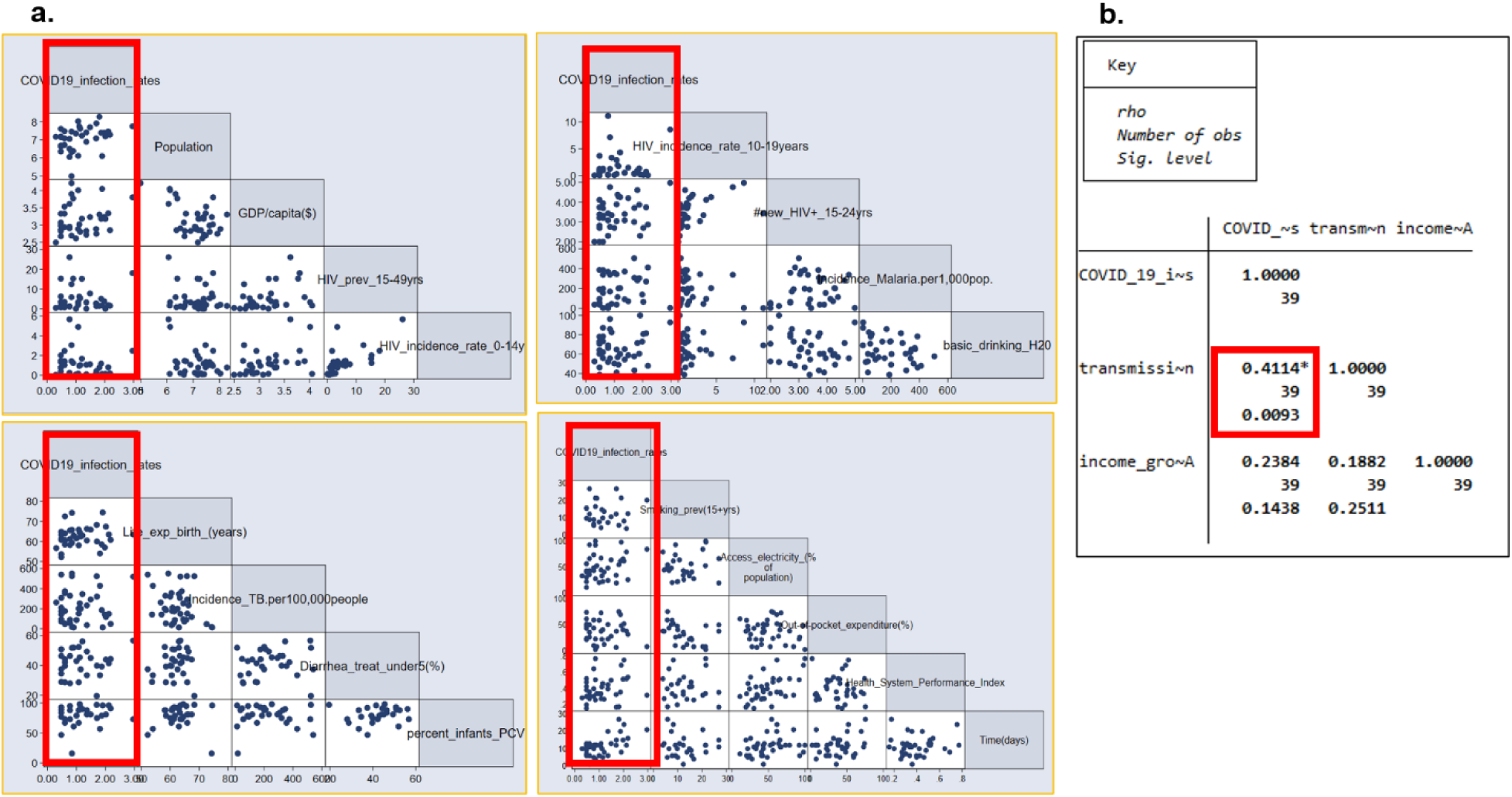
Summary Statistics of Continuous and Categorical Variables in the Machine Learning Model. (a. Matrix plot showing correlation of seventeen continuous variables and COVID-19 disease, (b. Spearman Rank Test of two categorical variables and COVID-19 disease.

We calculated time (days) between the first and last reported coronavirus cases by country and plotted a quadrant chart of confirmed COVID-19 cases against time. In Figure 3, majority, 32 (82%) of the countries reported imported transmission as the main way coronavirus is spreading *χ2* (1) = 16.03, *p*= 0.0001, while South Africa, Ghana, Senegal, Cameroon, Kenya, Rwanda and Liberia indicated local transmission as the source of COVID-19 transmission. There was a strong positive correlation between time (days) and COVID-19, (*r_s_* (37) = 0.599, *p* = 0.001), with time (days) explaining 36% of the variation in COVID-19. The result is suggestive that new cases of coronavirus tend to rise with rise in time (days).

**Figure 3:**
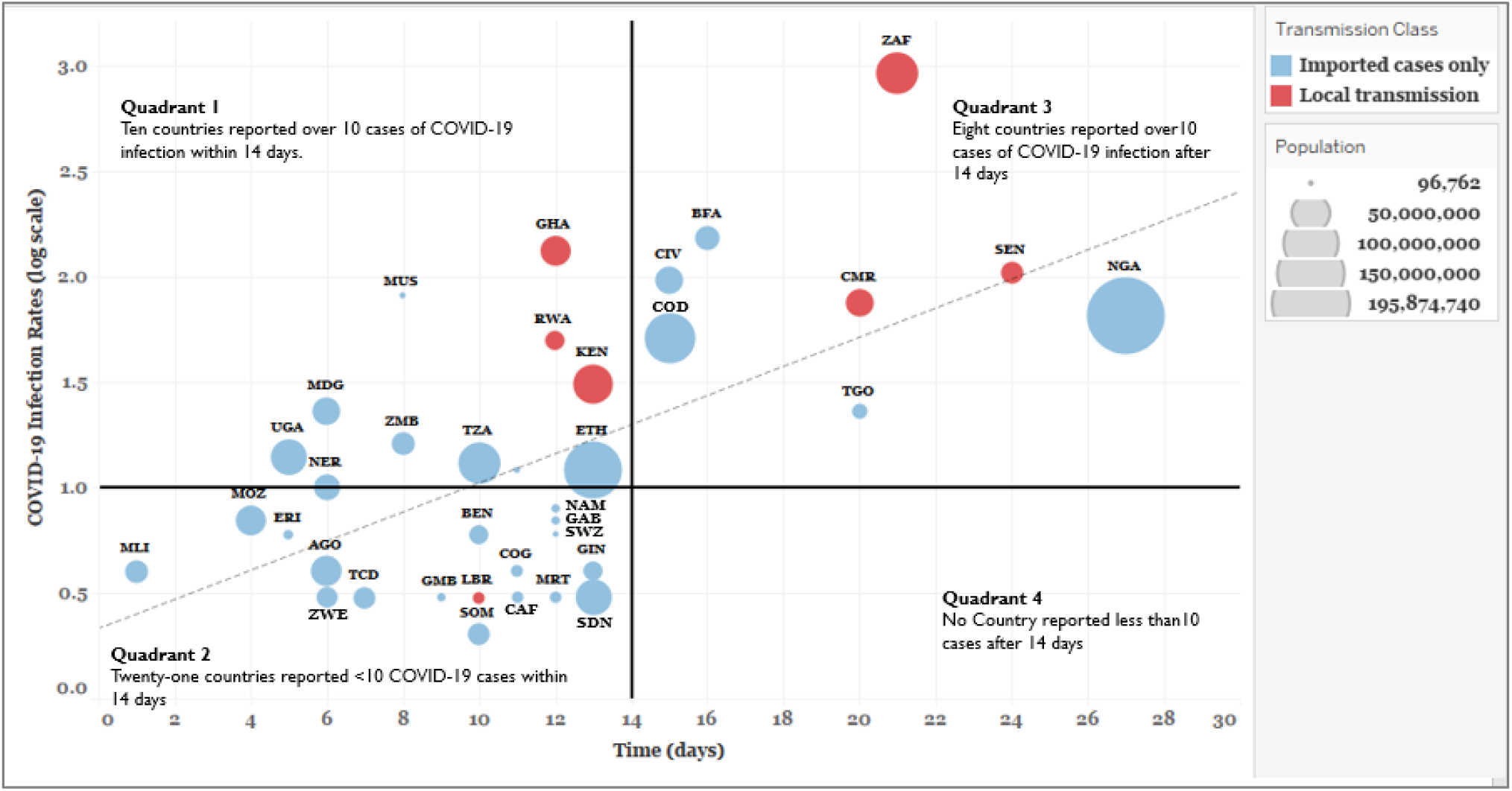
Status of COVID-19 Disease Outbreak in sub-Saharan Africa as of March 26, 2020.

Figure 4 shows steady and consistent growth in the total number of COVID-19 cases reported across SSA between March 21, 2020 to March 26, 2020. The steady decrease in the rate of change (ROC) of COVID-19 disease outbreak from 37% on March 22, 2020 to 23% on March 26, 2020 is suggestive of the impact of different aggressive measures by countries to stem the spread of coronavirus. We calculated daily ROC by dividing the current number of cases in each day by the number of cases from the earlier days and subtracted the value from 1.

**Figure 4:**
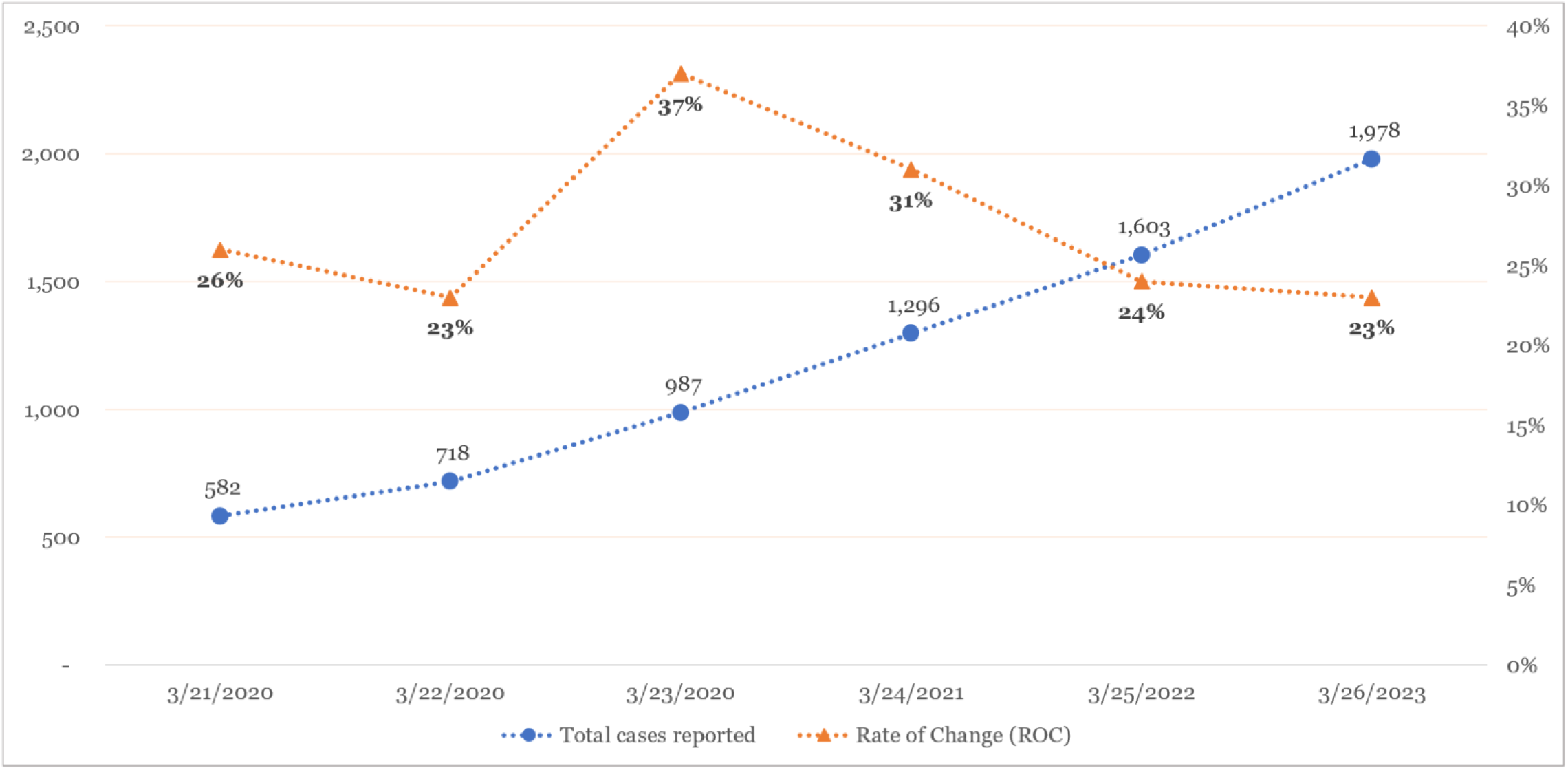
Trend and rate of change (ROC) of confirmed COVID-19 Cases in sub-Saharan Africa (March 21, 2020 - March 26, 2020)

A trend cluster analysis was performed using the cluster analytical tool in Tableau Desktop 2019 v.4 to determine if the increasing trends in new cases of coronavirus was similar across SSA countries. Five clusters were identified and South Africa the only country in cluster 5 (Figure 5). Overall, the result revealed an upward trend and similarities in the way coronavirus is presenting across certain countries in SSA.

**Figure 5:**
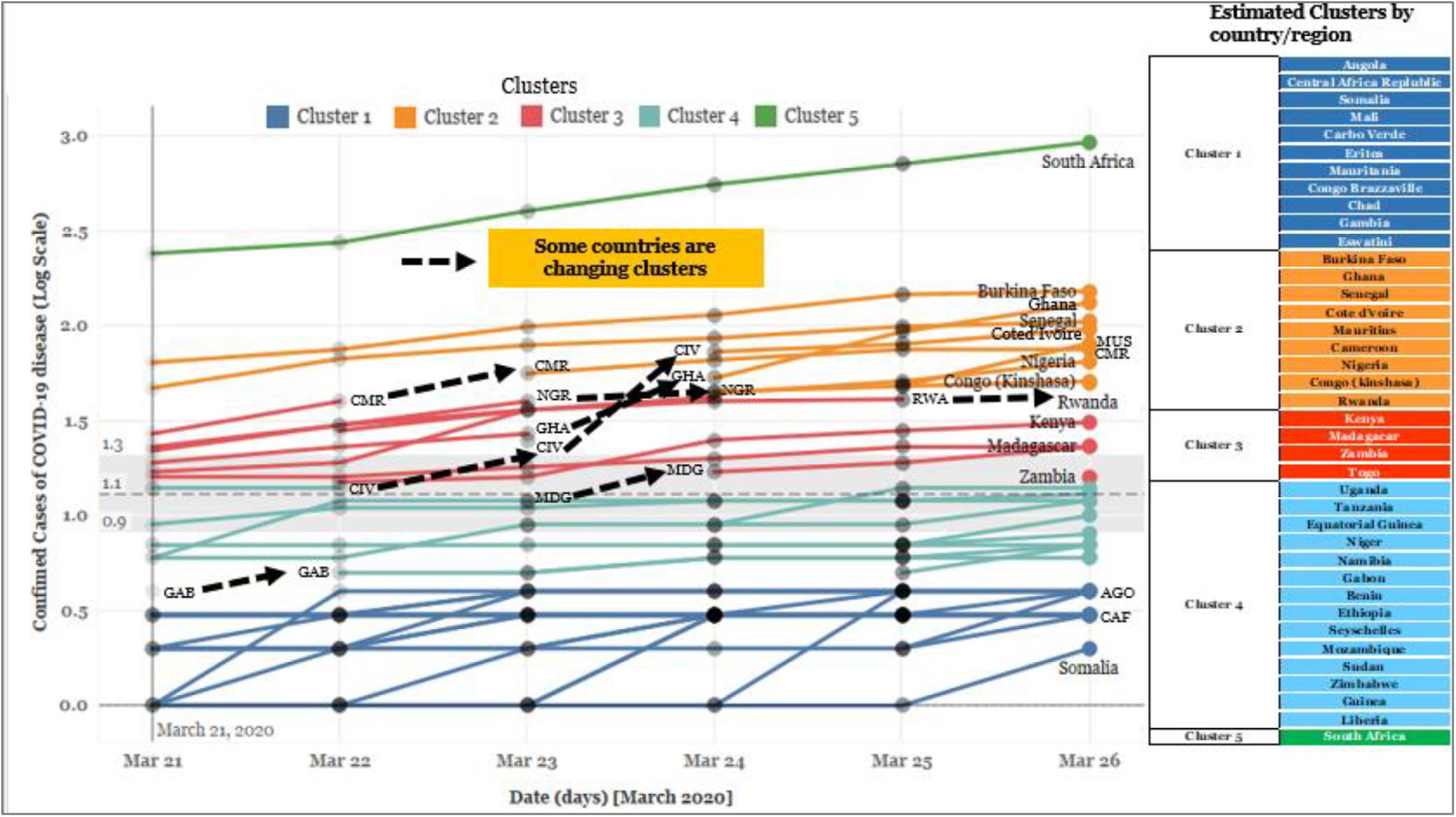
Coronavirus Trend Cluster Analysis (March 21, 2020 – March 26, 2020)

## Machine Learning

### Lasso for prediction and model selection

All the explanatory variables, 17 continuous and 2 categorical were included as possible contributors to the lasso model evaluating association with coronavirus disease. The rows for all three models were sorted so that the variables with the highest standardized coefficients are listed on top, (Table 1a.). Ten explanatory variables with the highest relative importance were simultaneously selected by all the three learning techniques. These were HIV incidence rate among adolescence aged 10–19 years, HIV incidence rate among children aged 0–14 years, Time (days), HIV prevalence among persons aged 15–49 years, infants who received third-dose of pneumococcal conjugate-based vaccine, Incidence of HIV among individuals aged 15–24 years, Incidence of malaria (per 1,000 persons at risk), Incidence of TB (per 100,000 people), Smoking prevalence (15years and above) and Diarrhea treatment (children under-5 years). We accessed the goodness-of-fit over our training sample and testing sample. (Table 1b.) show that the adaptive lasso model has the smallest mean square error (MSE), 2.72 *10^−28^ and the largest R-squared (100%) in the testing datasets.

**Table 1a):** Variables selected using three Lasso selection techniques. (The model displays an “x” if a variable was selected using any of the three methods, and most important variables are listed first.).

| Variables | cvplot | minBIC | adaptive |
| --- | --- | --- | --- |
| HIV incidence rate (persons aged 10-19 years) | x | x | x |
| HIV incidence rate (adolescence aged 0-14 years) | x | x | x |
| Time (days) | x | x | x |
| HIV prevalence (persons aged 15-49 years) | x | x | x |
| Percent infants who received PCV | x | x | x |
| Incidence of HIV (15-24 years) | x | x | x |
| Incidence of malaria (per 1,000 persons at risk) | x | x | x |
| Incidence of TB (per 100,000 people) | x | x | x |
| Smoking prevalence (15years and above) | x | x | x |
| Diarrhea treatment (children under-5 years) | x | x | x |
| Transmission classification = 1 |  | x | x |
| Transmission classification = 2 |  |  | x |
| Income group = 2 |  |  | x |
| Percentage of population with access to electricity |  |  | x |
| Income group = 3 |  |  | x |
| Life expectancy at birth |  |  | x |
| Population |  |  | x |
| GDP per capita |  |  | x |
| Health Systems Performance index |  |  | x |
| Percentage Out-of-pocket expenditure |  |  | x |
| Basic drinking water services |  |  | x |
| Income group =4 |  |  | x |

Legend: b - base level e - empty cell o - omitted x - estimated

**Table 1b):** Goodness of fit test.

| Name | Postselection Coefficients |  |  | obs |
| --- | --- | --- | --- | --- |
|  | Sample | MSE | R-Squared |  |
| cvplot | 1 | 0.1128382 | 0.752 | 11 |
|  | 2 | 0.0336583 | 0.9019 | 10 |
| minBIC | 1 | 0.0460822 | 0.8987 | 11 |
|  | 2 | 0.0530801 | 0.8452 | 10 |
| adaptive | 1 | 4.26e-28 | 1.0000 | 11 |
|  | 2 | 2.72e-28 | 1.0000 | 10 |

### Lasso for inferential statistics

The top ten important variables selected by adaptive lasso was included as variables of interest in the lasso for inference model using the Cross-fit partialing out technique. The remaining variables included in the model were trained as controls. The lasso regression for inference established that HIV incidence rate among adolescence aged 10–19 years (*p*=0.0001), HIV incidence rate among children aged 0–14 years (*p*=0.001), Time (*p*=0.0001), infants who received third dose of pneumococcal conjugate-based vaccine (*p*=0001), Incidence of HIV among individuals aged 15–24 years, (*p*=0.02), Incidence of malaria (*p*=0.001) and Diarrhea treatment (*p*=0.002) could statistically significantly predict coronavirus disease outbreak in SSA (Table 2). For better interpretation of results, we exponentiated the coefficients, subtracted the output from 1 and expressed the new coefficients as percentages. The exponentiated values are included in the lasso for inferential statistics table. All seven variables added statistically significantly to the prediction, *p* < 0.05.

**Table 2:**
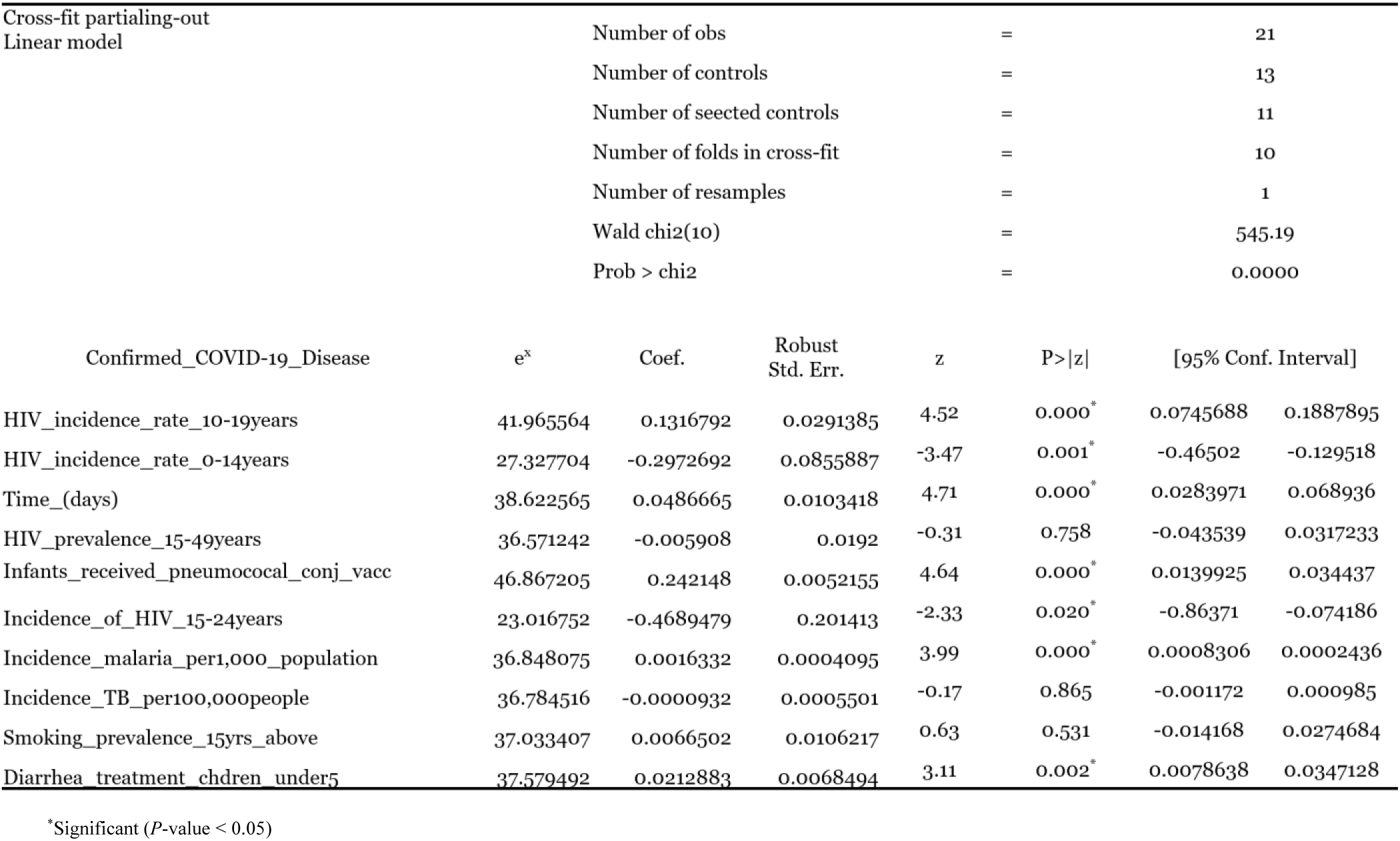
Lasso for Inferential Statistics: Cross-fit Partialing-Out Linear model.

### Geostatistical Analysis

The global spatial autocorrelation Moran’s I statistics confirmed the random distribution pattern of coronavirus outbreak among SSA countries (Figure 6). Given the z-scores of −0.63 (Moran’s I = −0.07, p=0.52), 0.22 (Moran’s I = −0.01, p=0.82) and −0.67 (Moran’s I = −0.04, p= 0.49) at March 8, 2020, March 14, 2020 and March 26, 2020 respectively, the pattern does not appear to be significantly differently from zero. EBK technique estimated the temporal spatial distribution of COVID-19 disease outbreak in SSA. The interpolated maps in Figure 7 shows that coronavirus is increasing per day and spreading outwards. Coronavirus appears to be severally confined in South Africa and increasing in the west African region (i.e. Burkina Faso, Ghana, Senegal, Cote d’Iviore, Cameroon, and Nigeria). Figure 8 shows that coronavirus deaths tend to be concentrated in western African regions relative to central and southern African regions.

**Figure 6:**
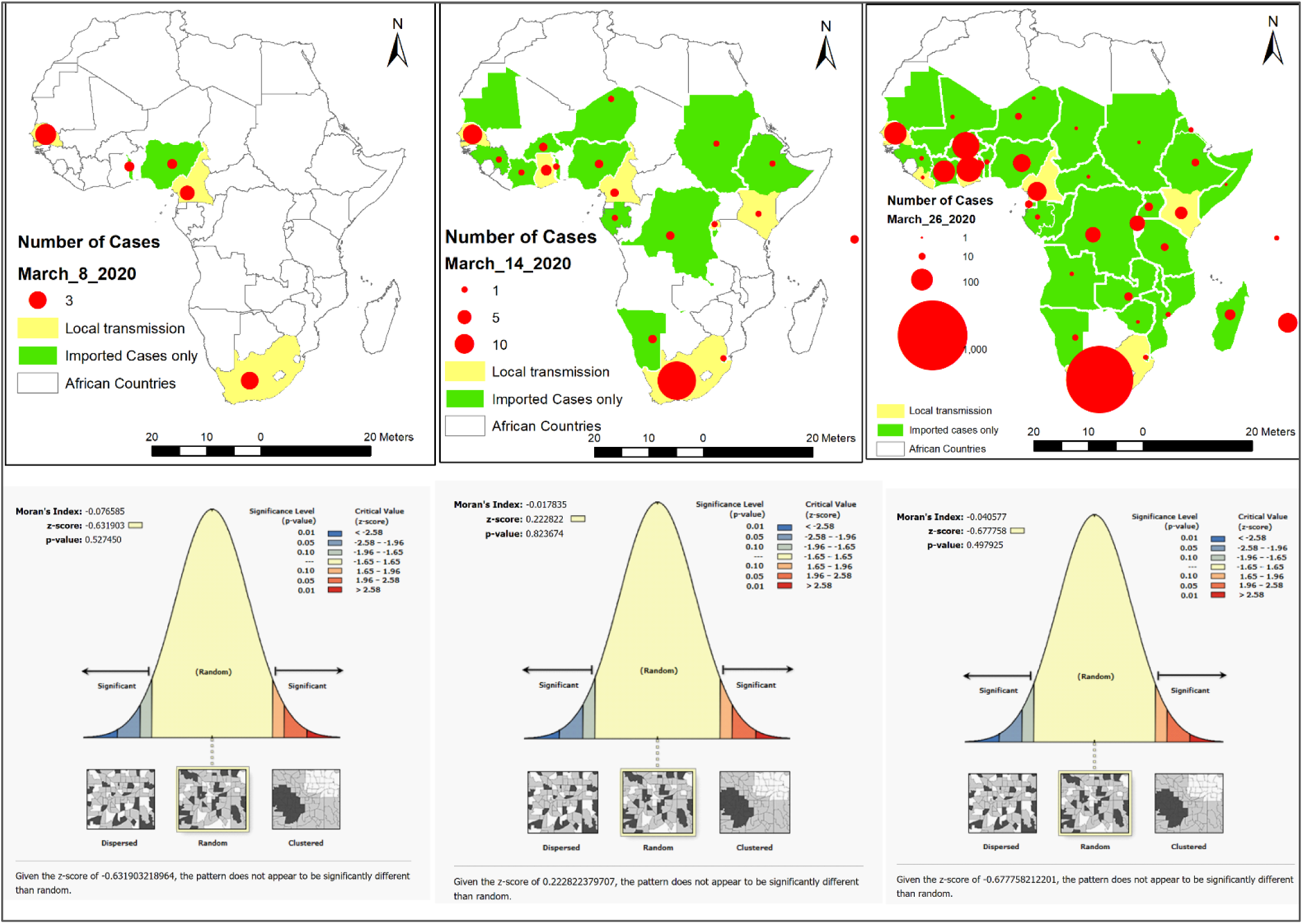
Spatial Autocorrelation analysis of COVID-19 Disease outbreak in SSA. (March 8, 2020, March 14, 2020 and March 26, 2020)

**Figure 7:**
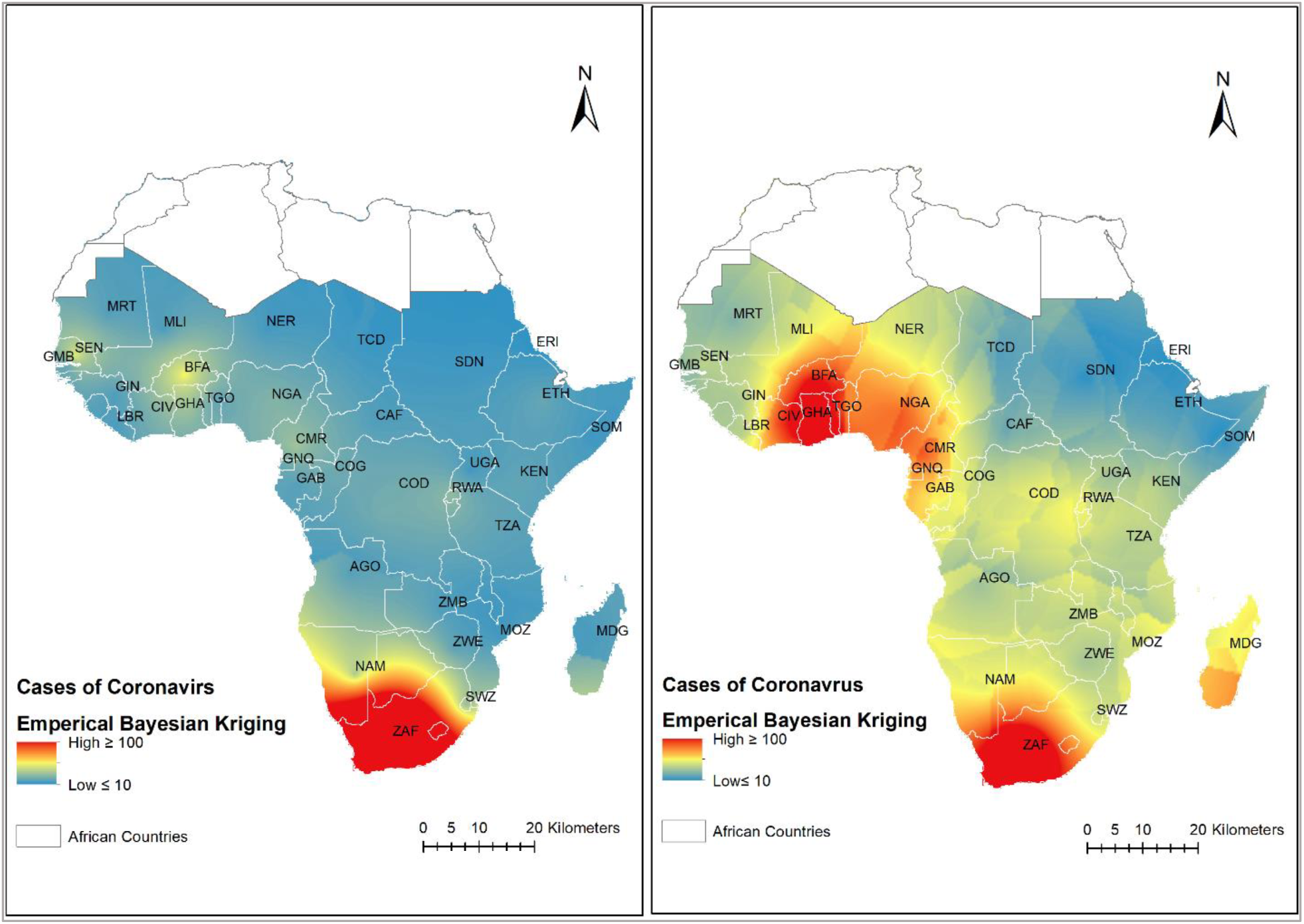
Interpolated Maps of COVID-19 Disease distribution across SSA using EBK technique. (a. Left panel – estimated distribution of COVID-19 disease at March 20, 2020 (b. Right panel – estimated distribution of COVID-19 disease at March 26, 2020)

**Figure 8:**
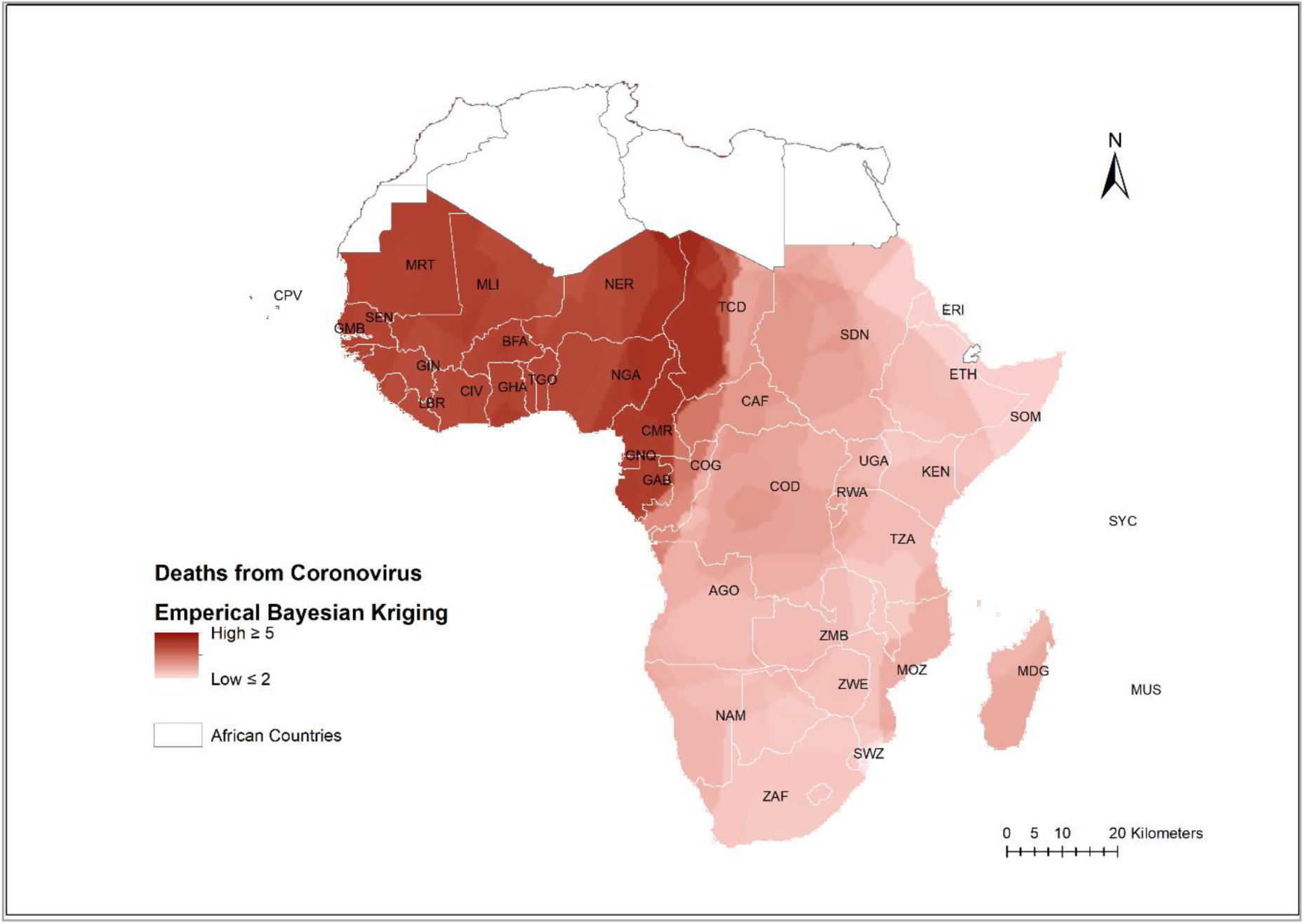
Interpolated Map of Deaths from COVID-19 Disease across SSA, March 26, 2020.

## Discussion

Of the estimated 6,000 new HIV infections that occur globally each day, two out of three are in sub-Saharan Africa^10^. Currently in SSA region there is paucity of research on coronavirus and no documented evidence on how coronavirus disease is affecting or behaving in people living with HIV (PLHIV). However, global and local HIV organizations and public health experts believe PLHIV not on treatment, has CD4 count <200 copies/cell, a recent opportunistic infection and /or not virally suppressed may be at greater risk of coronavirus infection^11^. Our study indicated that HIV incident rate among pediatrics aged 0–14 years, adolescents aged 15–19 years and middle-aged adults 15–24 years were good predictors of coronavirus. Our research shows that new HIV infections raise the risk of coronavirus infection by 27.3 percent (p=0.001) in pediatrics 0–14 years, 41.9 percent (p=0.0001) in adolescents aged 15–19 years and 23 percent (p=0.02) in middle-aged adults 15–24. This finding collaborates with a recent study by Su L et al^12^, which showed that children and youth are also infected and can spread coronavirus. A retrospective study by Tang A et al^13^ revealed that there had been several and far more severe pediatric cases of COVID-19 in Wuhan hospitals than reported.

Pneumonia affects children and families everywhere but is most prevalent in South Asia and sub-Saharan Africa^14^. People who get pneumonia may also have a condition called acute respiratory distress syndrome (ARDS). It’s a disease that comes on quickly after infection from coronavirus and causes breathing problems. The pneumonia vaccine protects against a kind of bacteria (Streptococcus pneumoniae), not the coronavirus, but can support overall health, especially in older individuals or people who have a weak immune system^15^. More than three-quarters, 79% (95% CI: 72.5–85.6) of surviving infants 12 to 23 months across SSA received the third dose of pneumococcal conjugate-containing vaccine (PCV). Our study indicates that surviving infants who received PCV were good predictors of coronavirus disease. It is evident from our results that increased cases of pneumonia among infants increases risk of infection from coronavirus by 46.8% (p=0.0001).

Malaria infection is common in Sub-Saharan Africa, and it is often stated that the continent bears over 90 percent of the global *P. falciparum* burden^16^, and, generally, young children bear the brunt of the mortality burden. In 2017, four SSA countries; Nigeria, Democratic Republic of the Congo, Mozambique, and Uganda accounted for nearly half of all malaria cases worldwide. According to our results, incidence of malaria was 189 per 1,000 persons (IQR:22 - 411) across SSA, and study results suggest that for every one-unit increase in the incidence of malaria, the risk of COVID-19 disease increases by 36.8% (p=0.0001).

Diarrheal disease is the second leading cause of death in children under five years old and is responsible for killing around 525,000 children every year. Recent studies have demonstrated that there is an association between diarrhea and coronavirus transmission. A study among 200 COVID-19 patients at three hospitals in Wuhan, China, indicated that among all patients who presented with digestive symptoms (117 patients), about 67 (58%) had diarrhea, and of these, 13 (20%) experienced diarrhea as the first symptom of their illness^17^. In our study, less than half 43% (95%CI: 28–56) of children under 5 with diarrhea received oral rehydration salts (ORT) or the recommended homemade fluids across SSA. Study result suggests that for every one-unit increase in diarrhea cases among children under-5, the risk of coronavirus increases by 37.5% (p=0.002).

In terms of growth rate, the trend cluster analysis from our study suggests a combination of linear and exponential growth trends across SSA. The trend in new coronavirus cases appears to be doubling in South Africa at every time increment compared to the other countries demonstrating an exponential trend. For countries in cluster 1 and in cluster 4 the trend is linear suggesting increases by less than 10 cases per day. Countries in cluster 2 and 3 on the average indicated increases by 10 cases or more at every time increment. Spontaneous transitions from low (reporting 10 cases on average) to high clusters (reporting more than 10 cases on average) was evident among countries previously in cluster 3 (i.e. Cote d’Iviore, Cameron, Nigeria and Ghana). In the coming weeks, our study suggests, that countries in cluster 2; Burkina Faso, Ghana, Senegal, Cote d’Iviore, Mauritius, Cameroon, Nigeria, Congo (Kinshasa) and Rwanda are the most likely to present an exponential trend growth similar to South Africa in cluster 5. Our study indicates, the doubling time in new coronavirus cases reported across SSA is 3 days.

COVID-19 WHO surveillance reports indicate that the SSA countries have responded actively to the pandemic since the first case identified on 28 February 2020. Many countries are requesting individuals to stay at home in self-quarantine, closing borders and imposing lockdowns and curfews. Schools were ordered to close in the Nigerian Federal Capital Territory of Abuja after only eight cases were confirmed nationwide in the month of February 2020. South Africa banned visitors from high-risk countries, closed schools and quickly opened drive-through testing centers in Johannesburg. The steady decline in the ROC of coronavirus as indicated in our study from 37% on March 23, 2020 to 23% on March 26, 2020 suggests that measures instituted by countries to stem the outbreak is showing signs of future success. This finding fits with a recent modeling study showing that a combination of multiple approaches (i.e. social distancing, home isolation of cases, augmented by school and university closures) has a substantial effect on slowing coronavirus transmission in a short term. On the other hand, the declining ROC may be attributed to low testing capacity in SSA. Marius Gilbert et al, in a modelling study highlighted differences in African countries capacity to respond to the coronavirus outbreak. Countries with the highest importation risk (ie, Egypt, Algeria, and South Africa) have moderate to high capacity to respond to outbreaks. Countries at moderate risk (ie, Nigeria, Ethiopia, Sudan, Angola, Tanzania, Ghana, and Kenya) have variable capacity and high vulnerability^18^. So far in SSA only a handful of countries (i.e. South Africa, Senegal, Ghana and Nigeria) have provided official figures on COVID-19 testing data.

Spatial Autocorrelation analysis using the Global Moran’s I statistics indicated that coronavirus was spreading randomly across SSA. A major limitation in this study was that geocoordinate data used for this analysis represents locations of SSA countries not necessarily where COVID-19 disease was detected, and this may have influenced results. At the time of this analysis, geocoordinates of counties, districts, and/or testing locations for coronavirus were not publicly available. The interpolated maps in our study show that coronavirus is increasing and spreading outwards per day to countries in central Africa and the virus appears to be severely confined in South Africa and in the west African region. The interpolated maps also suggest that countries in the west African region are most likely to repot increased number of deaths in the coming weeks compared to countries in central and Southern Africa.

## Conclusion

In conclusion, lasso was a good fit for variable selection and inference. The lasso regression model indicated that new HIV infections among pediatrics, adolescents and middle-aged adult PLHIV, time, pneumococcal conjugate-based vaccine, incidence of malaria and diarrhea treatment could significantly predict coronavirus disease in SSA. This study can be repeated using more detailed patient-level granular data to provide more insight into the essential characteristics of patients diagnosed with COVID-19. Our study suggests that experiences learned from the HIV epidemic can be applied to the fight against COVID-19. As in the AIDS response, governments should work with communities and civil society organizations to find local solutions, and health interventions delivered in an integrated manner. Integrated and efficiently delivered interventions to reduce HIV, pneumonia, malaria, and diarrhea are essential to accelerating global health efforts. Lessons learned from the HIV epidemics have shown that restrictive, stigmatizing and punitive measures can lead to significant human rights abuses, with disproportionate effects on already vulnerable communities. They can often undermine epidemic responses, sending people with symptoms underground and failing to address the underlying barriers that people face in attempting to protect their own health and that of their community. An approach that moves away from compulsory restrictions towards a focus on reaching and serving those who are most vulnerable, scaling up screening and testing for those most in need, empowering people with knowledge and tools to protect themselves and others (e.g. for COVID-19, increased social/spatial spacing) and the removal of barriers, mirrors the learnings from the HIV response.

The consistent three days decline in the ROC of coronavirus outbreak from 37% to 23% is suggestive of the positive effect of measures instituted by countries to stymie the outbreak. To gain better understanding on how the pandemic is progressing, and possibly ensure a sustained decline in the ROC of coronavirus outbreak, all countries should scale screening and increase their testing capacity and provide detailed and reliable testing data.

Recent non-randomized control studies have confirmed that hydroxychloroquine and azithromycin have been found to be efficient on SARS-CoV-2 and reported to be efficient in Chinese COV-19 patients. We propose a *silver bullet* approach to controlling the outbreak. This silver bullet approach here implies the use of presumptive malaria treatment, or the use of mass drug administration (MDA)– which have proved useful in some previous emergencies. Through MDA, all individuals in a targeted population will be given hydroxychloroquine and azithromycin – often at regular intervals – regardless of whether they exhibit symptoms of the disease.

## Data Availability

Data used for this analysis was obtained from the COVID-19 Data Resource Hub established by the Tableau community and included near real-time data compiled by Johns Hopkins University. Additional data from socio-demographic and health indicator surveys was derived from web resources of the World Bank, UNICEF, WHO and UNAIDS. All data used are publicly available, and sources are cited throughout.

https://github.com/CSSEGISandData/COVID-19/tree/master/csse_covid_19_data

## Contributors

AAO conceived of the study including design and method. He was the principal in data management, analyzed the data and drafted the article. AA, CO, AK, EJ, PG, GE, DO, DM and MR contributed to reviews of the draft and final version of the manuscript.

## Data sharing

All data used are publicly available, and sources are cited throughout.

## Acknowledgments

Data used for this analysis was obtained from the COVID-19 Data Resource Hub established by the Tableau community and included near real-time data compiled by Johns Hopkins University. Additional data from socio-demographic and health indicator surveys was derived from web resources of the World Bank, UNICEF, WHO and UNAIDS.

### Research in Context

#### Evidence before this study

The latest outbreak of the 2019 novel coronavirus disease (COVID-19) has spread rapidly within China and has spread worldwide afterwards. Certain epidemiological features and clinical characteristics of COVID-19 have been previously reported. However, these findings were based on prediction models or studies from America, Asia and Europe countries, and the use of these findings might be inappropriate for sub-Saharan Africa context. The spatial distribution pattern of coronavirus disease outbreak and the risk factors leading to poor clinical outcomes have not been well understood and delineated. We searched PubMed for articles in English published on and before March 26, 2020, that included “coronavirus”, “CoV”, “2019-nCoV”, “Machine Learning”, and “Empirical Bayesian Kriging”. Only two studies used predictive models of machine learning to either investigate the effect of COVID-19 on people’s mental health or forecast the occurrence of COVID-19.

#### Added Value of this Study

Given the scarcity of evidence of potential risk factors of COVID-19 in sub-Saharan Africa, we combined and analyzed multiple data streams using supervised machine learning algorithms, lasso to evaluate the importance of a set of nineteen explanatory variables to predict the risk of coronavirus infection. Through this, we identified seven variables that were significantly associated with the risk of coronavirus infection. We discussed the seven risk factors of coronavirus according to their relative importance as highlighted in our lasso model under two dimensions – 1.) infectious diseases: HIV, pneumonia, malaria, and diarrhea, and 2.) growth rate- time. To better understand the pattern and spatial distribution of coronavirus outbreak, we also used Empirical Bayesian Kriging to create a raster estimating the temporal spatial distribution of COVID-19 disease outbreak in SSA.

#### Implications of all the available evidence

The random distribution pattern of coronavirus infection as indicated in our study suggests that we cannot control the coronavirus pandemic by targeting interventions to a specific region or country. New cases and deaths from coronavirus are most likely to increase in the west African regions in the coming weeks. Managing the current COVID-19 pandemic requires a comprehensive and coordinated response within and between sub-Saharan countries and the continents, otherwise the pandemic will continue to surge at a greater cost to the global economy and public health.

